# Electrocorticography during deep brain stimulation surgery for movement disorders; Single-center experience

**DOI:** 10.1101/2025.07.13.25331460

**Authors:** Helena Ljulj, Kurt Lehner, Kimberley Wyse-Sookoo, Toren Arginteanu, Yousef Salimpour, William S. Anderson

**Author notes:** These authors contributed equally (Yousef Salimpour, William S. Anderson). **Correspondence:** Yousef Salimpour, MSc, PhD.

## Abstract

**Background:** Electrocorticography (ECoG) can be used as an intraoperative research tool during deep brain stimulation (DBS) implantation procedures. Its application has contributed to understanding the neurophysiology of movement disorders and the therapeutic effects of DBS. The aim of this report is to demonstrate the feasibility, safety, and utility of high-density ECoG for acquiring high-resolution neurophysiological data during DBS surgery.

**Methods:** Data were obtained from patients undergoing awake DBS surgery for the treatment of Parkinson’s disease (PD) or essential tremor (ET) at Johns Hopkins Hospital between February 2021 and September 2024. Burr holes created for the DBS lead implantation were used for ECoG strip placement. Electrophysiological and anatomical data were analyzed using MATLAB FieldTrip and Freesurfer, with localization in the anterior commissure and posterior commissure (ACPC) and Montreal Neurological Institute (MNI) coordinate systems. Surgical complications were monitored for at least six months postoperatively.

**Results:** Thirty-six patients (26 PD, 10 ET) were enrolled in the study. In one case, anatomical placement was inadequate for neurophysiological analysis. Postoperative complications included three infections (8.3%) and one chronic subdural hematoma (2.8%), with no permanent neurological deficits. The total complication rate was 11.1%, and all complications were unlikely to be related to ECoG strip placement. Anatomical and neurophysiological analysis demonstrated high-resolution cortical mapping. Multiple-subject level analysis using high-density ECoG yielded over 1,300 electrode positions.

**Conclusion:** ECoG during DBS is a valuable research method for movement disorders without additional risk to the standard procedure. The use of high-density intraoperative ECoG grids and the analysis of multiple-subject data in a standardized anatomical mapping space allows for high-resolution neurophysiological data acquisition and analysis.

## Introduction

Electrocorticography (ECoG) during deep brain stimulation (DBS) procedures presents a unique opportunity for intraoperative neuroscience research. DBS is a well-established procedure for movement disorders including Parkinson’s disease (PD) and essential tremor (ET), during which burr holes are made to allow access to deep brain targets. Through the same burr holes, subdural electrocorticography (ECoG) strip electrodes can be placed, allowing for localized cortical recordings.^1^ This makes it possible to obtain cortical ECoG signals while simultaneously collecting subcortical signals in the form of local field potentials (LFPs) and spiking neuronal activities. Because these procedures are conducted while the patient is awake and cooperative and recordings are obtained from both cortical and subcortical structures, real-time brain activity during behavior and stimulation can be captured.

Clinical application of ECoG is well-established in seizure detection and functional mapping of cortical areas during resective surgeries.^2,3^ Its use in research has been expanding, especially in fundamental neuroscience brain mapping,^4–6^ and more recently in the development of brain-computer and brain-machine interface systems.^7–10^An increasing number of studies use ECoG during DBS surgery to explore the cortical-basal ganglia network, helping to improve understanding of the brain mechanisms behind movement disorders such as Parkinson’s disease, essential tremor, and dystonia^11–18^. They were also essential in understanding the underlying mechanisms for the therapeutic effects of DBS in PD,^19^ and demonstrated the potential for advancements in the development of closed-loop stimulation systems for movement disorders such a PD and ET.^20–23^

Large-scale studies have confirmed the technical aspects, safety, and utility of conducting research using ECoG during DBS surgery. Two large multi-center studies have been particularly influential in establishing this. Sisterson et al. reported on a cohort of 200 patients, and Panov et al. on 367 patients undergoing DBS with concurrent ECoG recordings.^24,25^ Both studies demonstrated low complication rates, the technical feasibility, and the utility of the technique. However, most of these studies have been limited by a lower spatial resolution of the electrophysiological data as high-density grids suitable for intraoperative use were unavailable. Additionally, analyses were typically performed at the single-subject level; standardized pipelines for anatomical localization and multi-subject integration were not utilized and did not take advantage of normalized coordinate systems to align electrode positions across subjects, limiting the ability to perform high-resolution group-level analyses.

Recent advancements, both in electrode technology and in anatomical mapping methods, have addressed these limitations and now enable higher resolution ECoG research during DBS. This report, presents a series of 36 patients undergoing awake DBS surgery for PD or ET, during which high-density ECoG strips were placed through the burr holes. The report aims to demonstrate the feasibility, safety, and technical consistency of the approach and presents both anatomical and neurophysiological data at the single and group subject levels. This work supports the growing role of intraoperative ECoG in studying movement disorders and contributes to the foundation for future research into adaptive, closed-loop neuromodulation systems.

## Methods

### Patient selection and consent

Patients considered for participation in the study were scheduled for DBS lead placement surgery for PD or ET in the “awake” state (light IV dexmedetomidine sedation was used in most cases), at the Johns Hopkins School of Medicine between February 2021 and September 2024. The exclusion criteria included inability to undergo awake DBS and any other exclusion as surmised by the multidisciplinary movement disorder surgery committee at Johns Hopkins (i.e. significant subcortical dementia, or other surgically prohibitive co-morbidity). The study was approved by the Johns Hopkins Institutional Review Board. Patients were informed about the study during an in-person meeting with the neurosurgeon, and written informed consent was obtained.

### Surgical planning

A preoperative MRI was obtained and imported into the DBS planning system (Brainlab Elements, Brainlab AG, Munich, Germany). Based on the condition (PD, ET) the subcortical target (the subthalamic nucleus (STN) or ventral intermediate nucleus (VIm) of the thalamus, respectively) was selected. The entry point of the lead and the burr hole location was chosen solely based on clinical criteria, with consideration to avoid large vasculature, sulci, and the ventricular system. The ECoG target cortical coverage was marked on the cortical surface using the planning system. The cortical target area for PD and ET was the ‘hand knob’ of the primary motor cortex area inferred from the location of the precentral gyrus.

### ECoG placement

All surgical procedures were performed with robotic stereotactic assistance (ROSA, Zimmer Biomet, Warsaw, IN or Globus ExcelsiusGPS, Globus Medical, Audubon, PA). A Leksell stereotactic frame was placed on the patient’s head and used for head fixation during the surgery. After positioning, a 3D cone beam intraoperative CT image set was obtained (Medtronic O-arm, Medtronic, Minneapolis, MN). This data was downloaded onto the robotic system and co-registered with a previously acquired head CT and MRI examination.

Following the robotic system registration process, the scalp was marked at the sites for the anticipated burr holes based on the DBS implant trajectory. The targeted cortical areas for ECoG strip placement were marked on the scalp. After skin incision and scalp dissection, a perforator was used to drill bilateral burr holes. The burr hole lead fixation system rings were then securely mounted. All patients underwent unilateral ECoG recordings based on the site of the initial DBS lead implant, which was typically the patient’s most symptomatic side. The dura was opened widely with a scalpel, and the dural leaflets were removed to prevent potential deflection of the DBS leads. The brain was gently depressed with a Penfield #3 spatula, and the subdural recording strip electrode was passed into the subdural space, aiming to center the electrode under the marked cortical site. The cabling was secured to the drapes with a clamp. The strip electrode cabling was then connected via the appropriate connection cables to the data acquisition system (Blackrock NSP, Blackrock Microsystems, Salt Lake City, UT).

The insertion guide tube cannulae for microelectrode recordings and DBS electrode placement were inserted through the burr hole and the working arm of the robotic system, followed by the microelectrodes (Alpha-Omega NeuroProbe, Alpha-Omega Engineering, Nazareth Illit, IS). Following placement of the microelectrodes, the burr hole was covered with gelfoam and fibrin glue to prevent CSF egress and minimize brain shift during the experimental recording sessions. Stimulation testing using the mapping microelectrode system and stimulation through the DBS leads was used to localize the optimal target for lead placement. Following localization of the DBS target with the microelectrodes, the patient performed the experimental tasks, which took no more than 25 minutes.

The placement of the subdural strip and microelectrodes was verified using the fluoroscopic mode of the O-arm and a 3D CT O-arm spin (Figure 1). Following this, the subdural strip electrode was removed and the DBS lead was placed and positioning was confirmed with macrostimulation mapping. Within 24h of the surgery, a postoperative CT or MRI was obtained as part of routine care.

**Figure 1.**
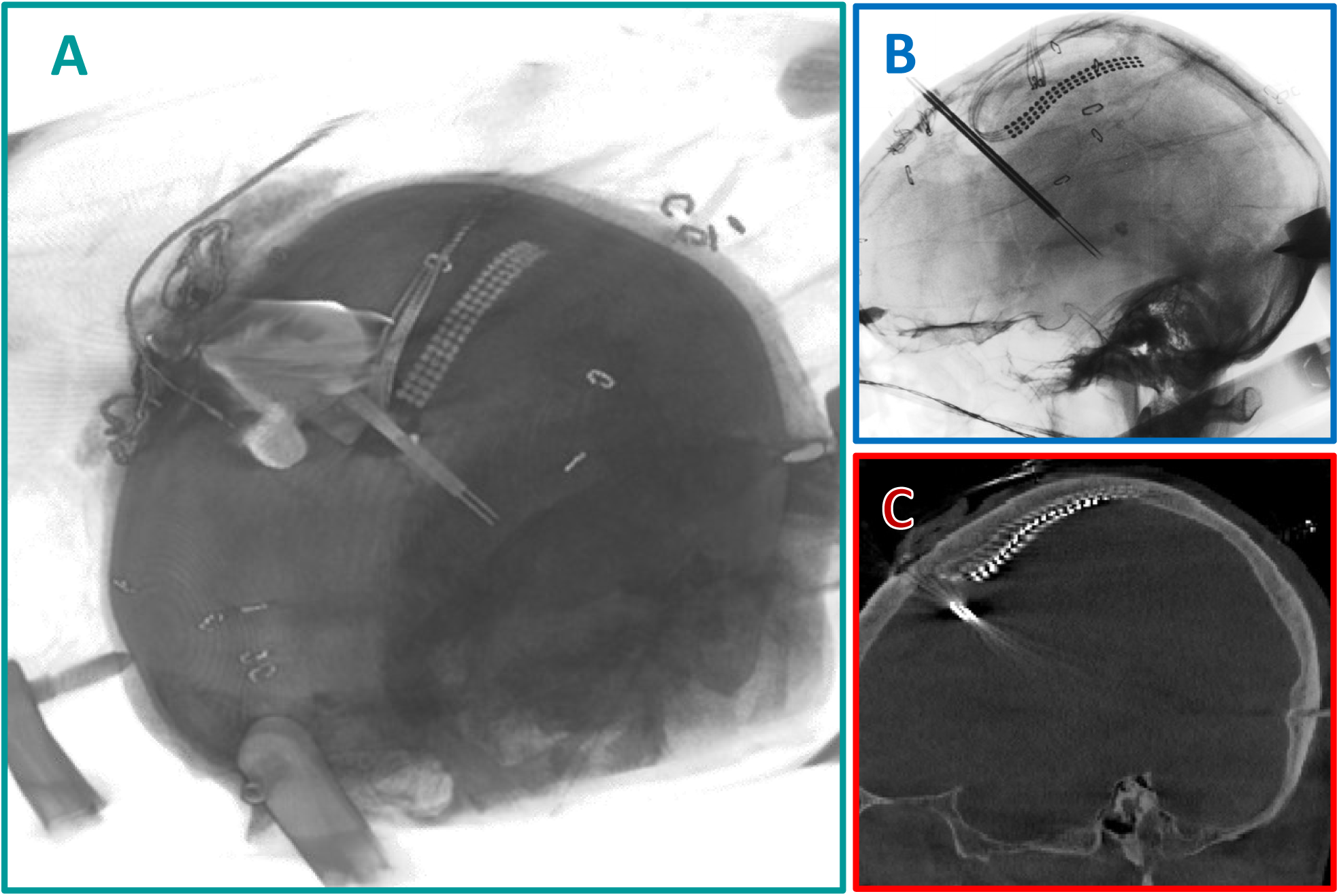
Intraoperative imaging showing the subdural placement of a 63-contact ECoG strip and the DBS leads at their target. (Upper) Left-sided lateral fluoroscopy image (Lower), intraoperative CT (O-arm, Medtronic) image.

### ECoG characteristics

Subdural strip electrodes (PMT Corporation, Chanhassen, MN, USA) used for electrocorticography (ECoG) contained 63 numbered contacts arranged in a 13 × 5 grid configuration (13 rows, 5 columns). Each circular contact had a diameter of 1 mm with 3 mm center-to-center spacing. The overall electrode strip measured approximately 6.85 cm in length and 0.9 cm in width, with a thickness of 0.76 mm. Electrodes featured inline tail connections, 4 leads, and color-coded tails (blue, green, violet, white) for ease of channel identification.

### Analysis of anatomical and electrophysiological data

Analysis of anatomical and electrophysiological data on single and multiple subject levels was performed following the protocol by Stolk et al.^26^ Using the FieldTrip toolbox^27^ for MATLAB, preoperative T1-weighted MRI scans and postoperative CT scans were co-registered and electrode coordinates were aligned. This allowed for anatomical localization of the contacts in the anterior commissure posterior commissure (ACPC) coordinate system.

Brain surface reconstruction using Freesurfer (version 7.4.1, http://surfer.nmr.mgh.harvard.edu/) was performed based on the preoperative MRI. This allowed visualization of the subdural strip contacts in the ACPC coordinate system on the native brain surface anatomy. Figure 2 displays the sample cortical structures covered by strip electrodes, including the motor cortex, dorsolateral prefrontal cortex, superior temporal gyrus, and posterior parietal cortex. Following anatomical analysis, the neurophysiological data were processed and visualized on the exact reconstruction.

**Figure 2.**
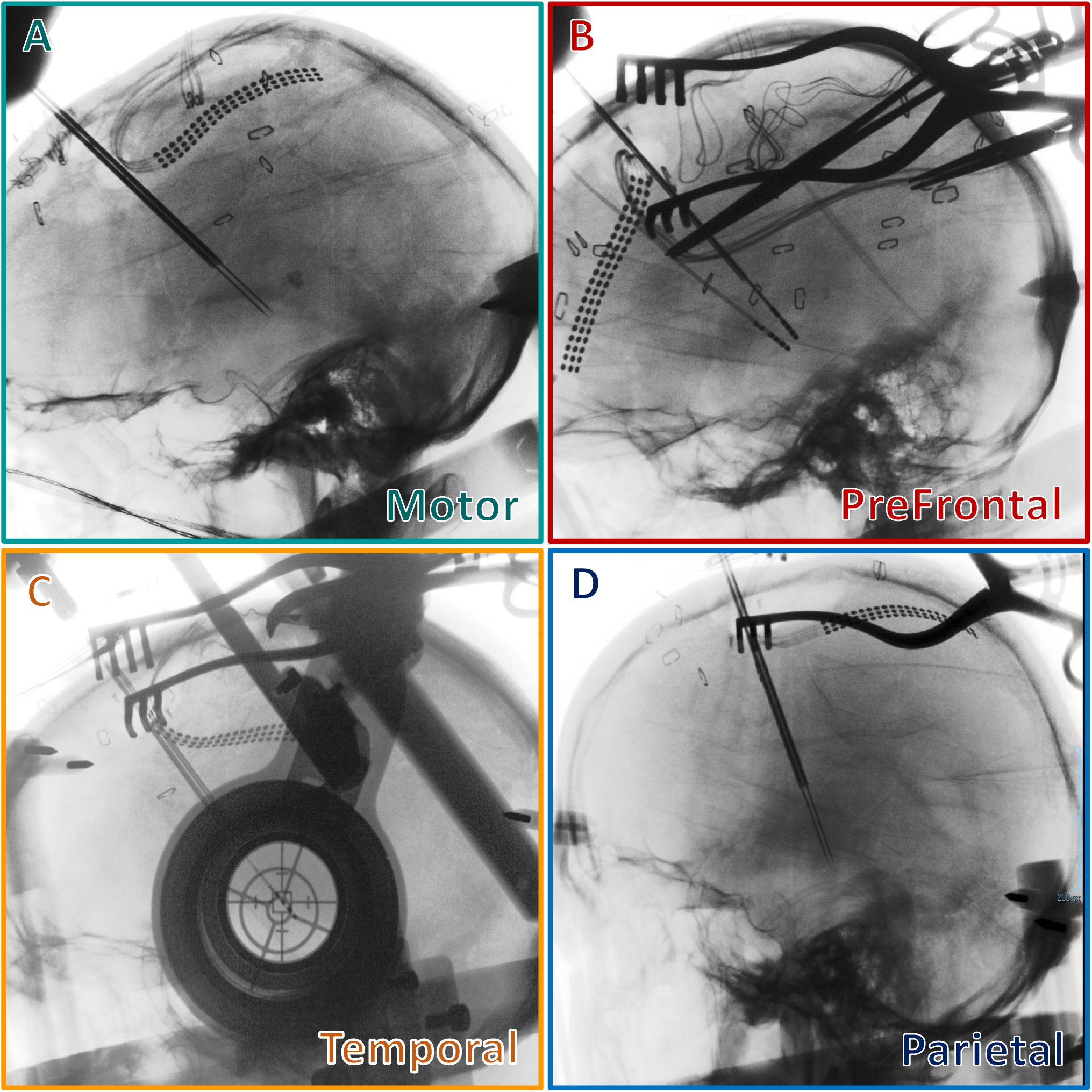
Fluoroscopic intraoperative images are displaying the subdural placement of a 63-contact ECoG strip while targeting various structures, including A. Motor cortex, B. Dorsolateral prefrontal cortex, C. Superior temporal gyrus, and D. Posterior parietal cortex.

For the purpose of multiple subject neurophysiological analysis, anatomical data of multiple subjects was integrated on a single normalized brain. ACPC contact coordinates for each subject were transformed to the Montreal Neurological Institute (MNI) coordinate space.^26^ This was performed using a non-linear volume-based normalization procedure implemented in SPM12 (https://www.fil.ion.ucl.ac.uk/spm/), aligning individual anatomical MRI scans to the MNI template, after which contact coordinates were transformed accordingly. MNI contact coordinates for each subject were then plotted on a standardized MNI brain template, allowing for data integration.

### Patient data and surgical complications

Routine postoperative CT scans, taken within the first 24 hours after surgery, were examined for possible hemorrhage, infarcts, or other complications. Patients were monitored for at least 6 months postoperatively, and surgical complications (such as infections, chronic subdural hematoma, intraparenchymal hematoma, or venous infarction) were recorded in a database.

## Results

### Patient demographics and ECoG anatomical coverage

A total of 36 patients were included in the study, with 26 undergoing DBS implantation for Parkinson’s disease (PD) and 10 for essential tremor (ET) (Table 1). In 34 cases, subdural ECoG strips were placed over the sensorimotor cortex using the same burr hole created for DBS electrode placement. In two patients, the strips were targeted to the dorsolateral prefrontal cortex for research purposes unrelated to motor physiology. In one case, strip placement did not result in adequate coverage of the primary motor cortex, specifically the hand knob area, and was excluded from group-level physiological analysis due to a lack of usable physiological signal.

**Table 1:**
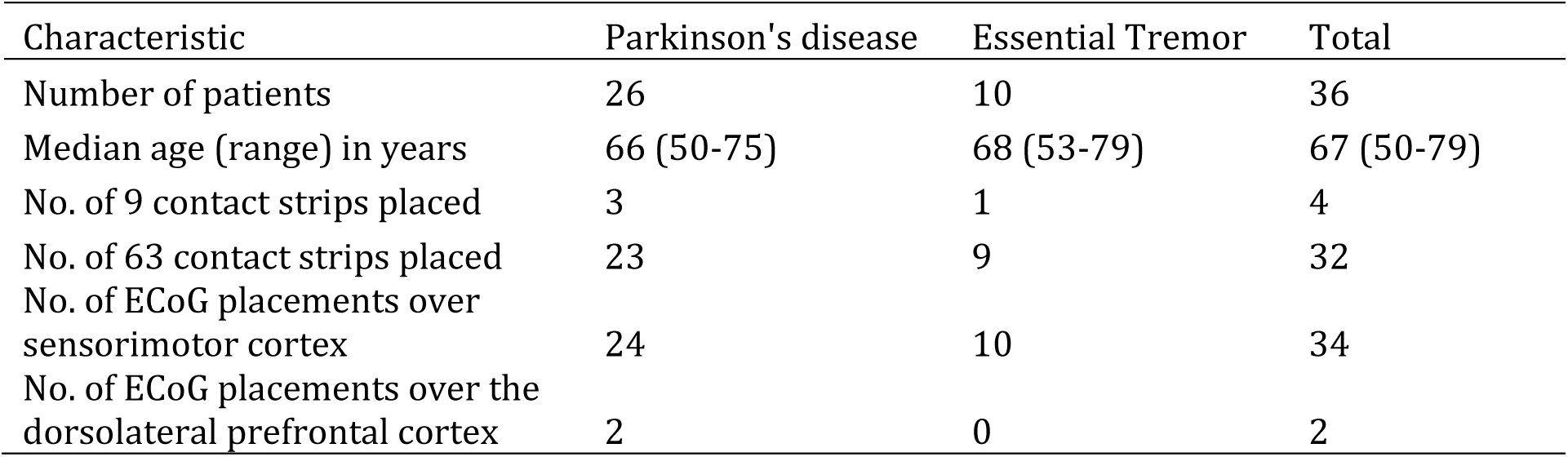
Patient demographics and ECoG anatomical coverage.

| Characteristic | Parkinson's disease | Essential Tremor | Total |
| --- | --- | --- | --- |
| Number of patients | 26 | 10 | 36 |
| Median age (range) in years | 66 (50-75) | 68 (53-79) | 67 (50-79) |
| No. of 9 contact strips placed | 3 | 1 | 4 |
| No. of 63 contact strips placed | 23 | 9 | 32 |
| No. of ECoG placements over sensorimotor cortex | 24 | 10 | 34 |
| No. of ECoG placements over the dorsolateral prefrontal cortex | 2 | 0 | 2 |

### Safety and complications

A total of four postoperative complications were observed, resulting in an overall surgical complication rate of 11.1% (Table 2). Three patients developed infections (8.3%), while one patient experienced a chronic subdural hematoma (2.8%). Of the infections, one was at the burr hole incision, two were at the superoauricular incision, and one occurred at the pulse generator site. The burr hole incision infection occurred ipsilateral to the side of ECoG strip placement. Importantly, the timing and location of these infections did not indicate a direct link to the ECoG strip.

**Table 2.** Postoperative complications.

| Complication | No. of cases | Reported rate in the literature |
| --- | --- | --- |
| Infection | 3 (8.3%) | Up to 9.9%<sup>28,29</sup> |
| Subdural hematoma | 1 (2.8%) | 3.0%<sup>28</sup> |
| Intraparenchymal hematoma | 0 (0.0%) | 0.6-2.9%<sup>28,29</sup> |
| Venous infarction | 0 (0.0%) | 0.8%<sup>30</sup> |

The patient who developed a subdural hematoma presented with worsening headaches two months after surgery. Imaging confirmed a chronic hemorrhage on the hemisphere contralateral to the ECoG strip, and surgical drainage was performed without complication. At the latest follow-up, this patient had no lasting neurological deficits.

### Example of processing anatomical and physiological data

Anatomical and neurophysiological data were analyzed at both the single-subject and group-subject levels. Electrode contact positions were localized using a combination of preoperative MRI and postoperative CT imaging, co-registered in ACPC space and transformed into MNI coordinates for cross-subject analysis. This allowed for precise anatomical mapping of each ECoG strip on the cortical surface and the integration of electrode positions across patients.

Figure 3 illustrates a representative single-subject analysis. Electrode positions are shown on both the subject’s native brain surface in ACPC space (Figure 3A) and on a normalized brain in MNI space (Figure 3B). Corresponding neurophysiological data, resting-state beta power, are visualized in Figure 3C, demonstrating spatial variability in cortical activity across the grid.

**Figure 3.**
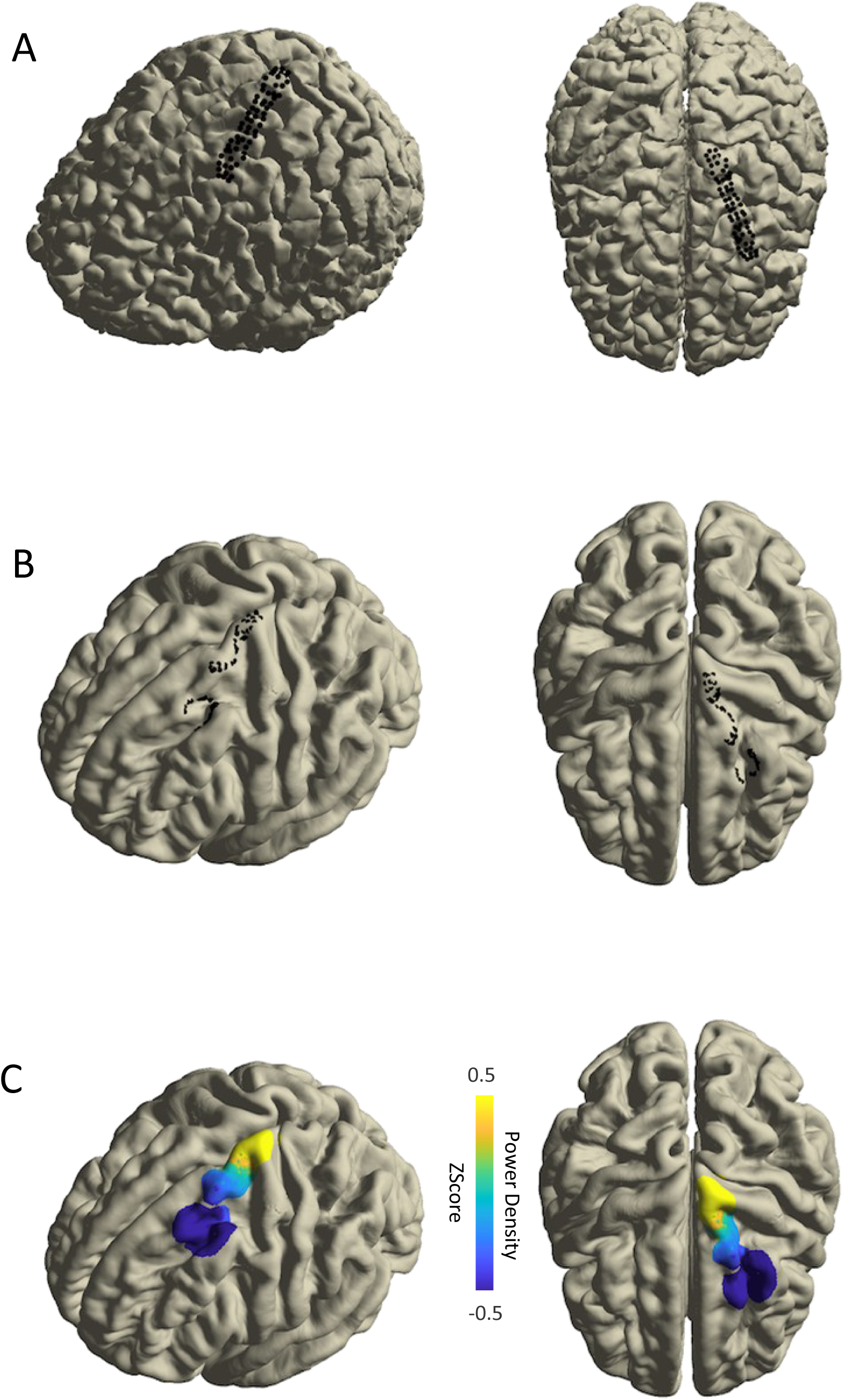
Single-subject analysis of anatomical and neurophysiological data. (A) Contact visualization on the native brain in the anterior and posterior commissures coordinate system. (B) Normalized contact positions visualization on an MNI brain reconstruction. (C) Neurophysiological data (beta frequency band power at rest) mapped onto the MNI brain.

Group-level anatomical analysis was performed on a subset of 19 patients with STN-targeted implantation for PD. This included eight patients with left-sided and eleven with right-sided ECoG strips, each consisting of 63 contacts. Electrode positions from all subjects were projected onto a standardized MNI brain to generate a group anatomical map (Figure 4), allowing for the visualization of consistent sensorimotor coverage across the cohort.

**Figure 4.**
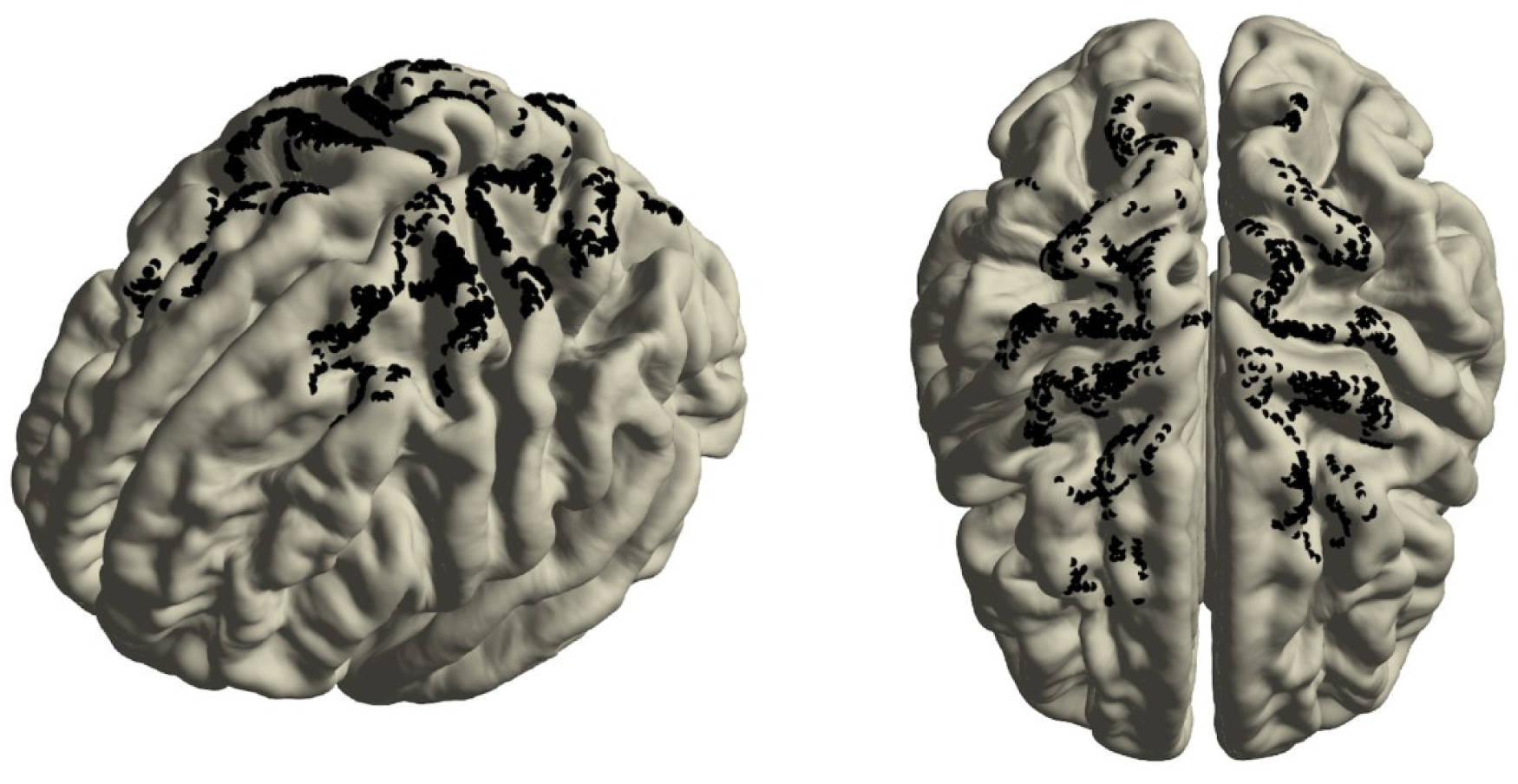
Multiple-subject anatomical analysis. MNI-space electrode localization on a normalized MNI space brain for 21 subjects.

Figure 4 shows the integration of group-level anatomical and physiological data. Resting-state beta activity is displayed across multiple subjects on a common cortical surface, highlighting both the feasibility and utility of high-density ECoG in capturing detailed spatial patterns of oscillatory activity during DBS procedures. To illustrate the combination of anatomical localization and electrophysiological measurements, Figure 5 displays the locations of electrodes across the subjects in a normalized brain model. It also presents the activity in the beta frequency range, which is used to map the motor cortex.

**Figure 5.**
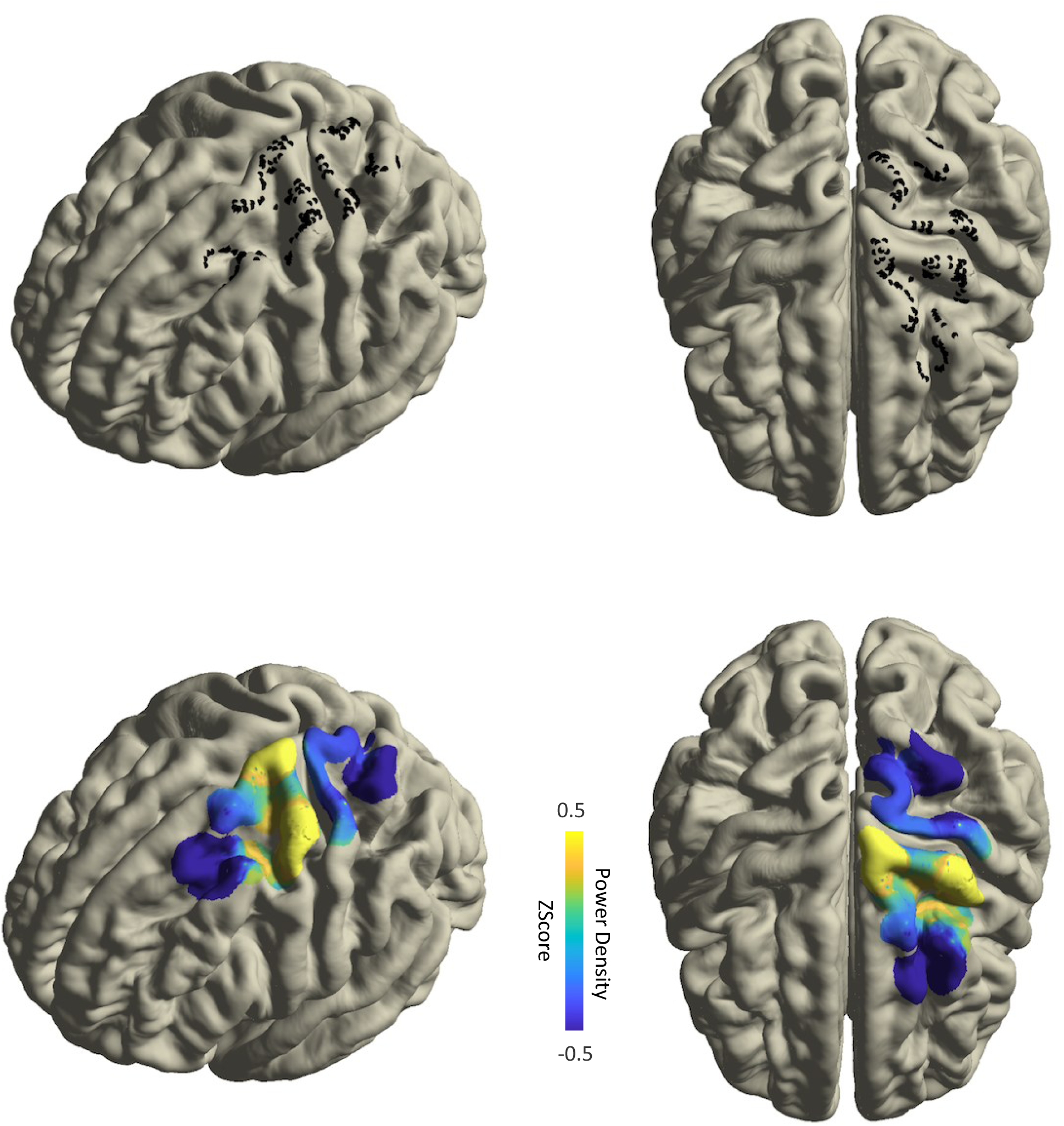
Multiple-subject analysis of anatomical and neurophysiological data. (Upper) MNI-space electrode localization on a normalized MNI space brain for three subjects. (Lower) Neurophysiological data (beta frequency band power at rest) mapped onto the MNI brain for three subjects.

After the electrodes are implanted in the designated target area, various intraoperative paradigms can be utilized during the surgical procedure of deep brain stimulation lead implantation. Taking advantage of this opportunity provides unique access to the human brain, allowing for the collection of valuable neurophysiological data. Figure 6 shows electrophysiological measurements from the motor cortex of the representative subject. The persistence spectrum of beta band power, which provides a time-frequency representation of the percentage of time that beta oscillations are present in the motor cortex of a Parkinson’s disease (PD) patient during intraoperative recordings at rest, is illustrated in Figure 6. A. The figure shows that as the duration of beta oscillations in the motor cortex increases over time, the percentage of time they are present rises, resulting in a brighter or “hotter” appearance in the display. The phase-amplitude coupling of the ECoG signal from panel A is clearly demonstrated using the modulation index quantification method (Figure 6.B). Mapping of the 63-contact ECoG strip electrode to the cortical surface is presented here, using anatomical information, beta band power, and beta-gamma phase-amplitude coupling measurements.

**Figure 6.**
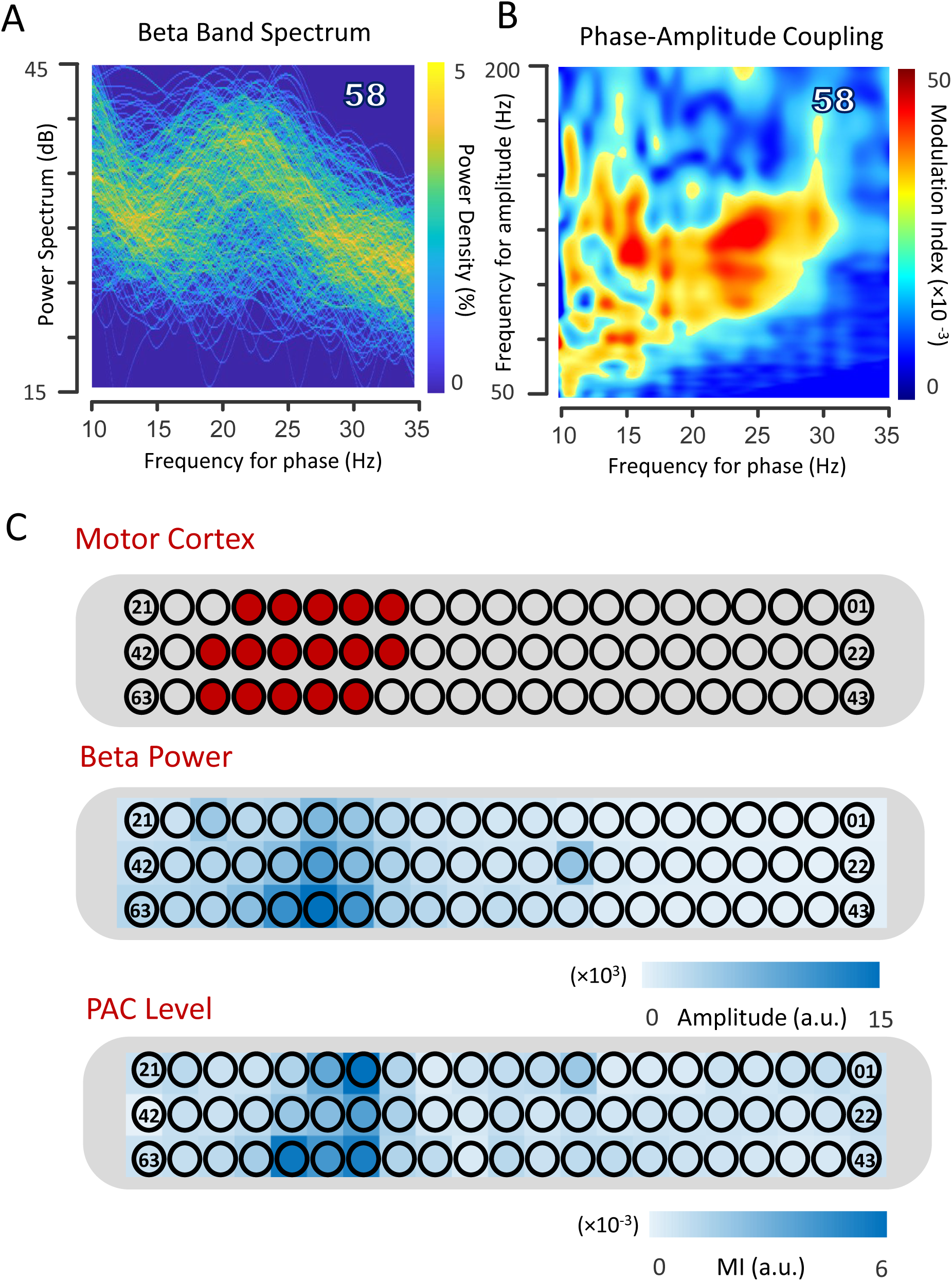
A. The persistence spectrum of the beta band power at rest, which is a time-frequency view showing the percentage of time that beta oscillations are present in a motor cortex recording, is displayed. B. Phase-amplitude coupling for the same ECoG signal from panel A is shown using the modulation index quantification method. C. Mapping of the 63-contact ECoG strip electrode to the surface of the cortex is shown here, utilizing anatomical information, beta band power, and beta-gamma phase-amplitude coupling measurements.

In addition to passive intraoperative recording, a variety of cognitive and motor-related paradigms can be conducted during surgical procedures. Figure 7 illustrates beta band dynamics during an upper limb motor task. The top section shows the beta band filtered signal, while the bottom section presents time-frequency analysis recorded during a repetitive grasping task from a subject with PD. Green lines indicate when the subject received a visual cue to start moving, and red lines indicate when the subject received a visual cue to stop moving.

**Figure 7.**
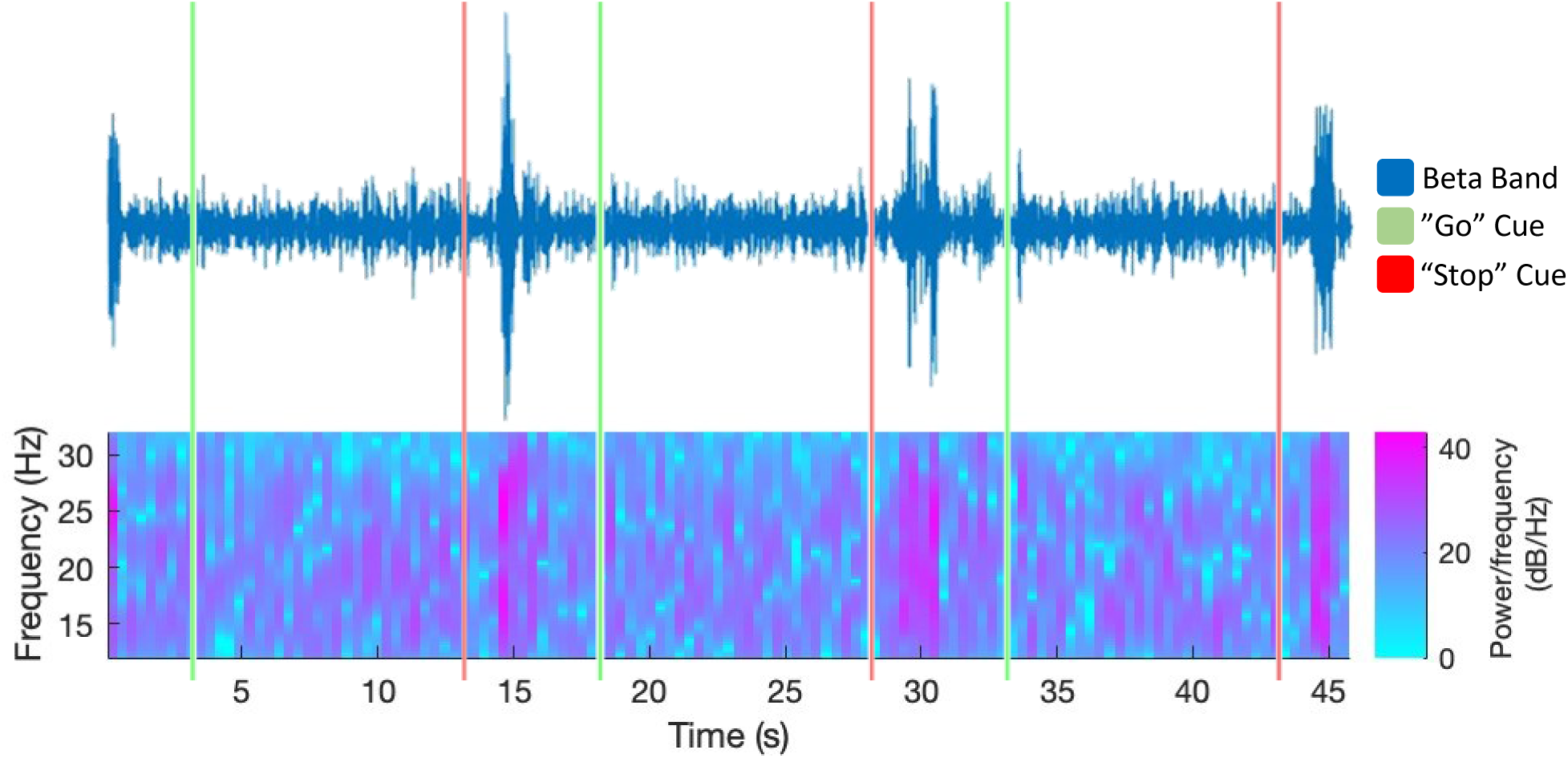
Beta band dynamics during an upper limb motor task. The beta band filtered signal (top) and time-frequency analysis (bottom) recorded during a repetitive grasping task from a subject with PD are shown. Green lines indicate when the subject was shown a visual cue to start moving, and red lines indicate when the subject was shown a visual cue to stop moving.

## Discussion

ECoG performed during DBS procedures can be a valuable tool for neurophysiological research on movement disorders. This report demonstrates the methodology, safety, and utility of the technique, highlighting how recent advancements in electrode technology and standardization in anatomical mapping across multiple subjects have enabled higher-resolution neurophysiological data acquisition and analysis.

### Safety and complications

As ECoG strip placement during DBS procedures is performed solely for research purposes and provides no direct therapeutic benefit, ensuring patient safety is essential. In this cohort, the overall complication rate was 11.1%, with three postoperative infections (8.3%) and one chronic subdural hematoma (2.8%). In comparison to a standard DBS procedure, the complication rate was lower for each of the complications, including infections,^28,29^ subdural hemorages,^30^ intraparenchymal hemorages,^30,31^ and venous infarctions.^32^ Notably, none of the complications were clearly attributable to the ECoG strip itself. One infection occurred at the burr hole incision site ipsilateral to the strip placement, but it presented one month postoperatively, making a direct causal relationship unlikely. The chronic subdural hematoma developed on the hemisphere contralateral to the ECoG strip and was successfully treated with surgical drainage, with no lasting neurological deficits.

These findings are consistent with previously published large-scale studies. Sisterson et al. ^25^ (200 surgeries) and Panov et al. ^24^ (367 surgeries) reported similarly low complication rates for DBS procedures incorporating ECoG, including infections, chronic subdural hematomas, intraparenchymal hemorrhages, and venous infarctions. These complication rates were equal to or lower than those observed in standard DBS surgeries without ECoG. Our findings reinforce the safety of incorporating subdural ECoG into DBS workflows, particularly when electrodes are inserted through existing burr holes.

As the aim of a large number of studies focusing on ECoG during DBS is ultimately the development of implantable, closed-loop neuromodulation systems, it is important to emphasize that results in this report apply specifically to the short-term intraoperative setting. Further investigation and studies are needed for long-term implantation of ECoG strips and grids. Existing literature on extraoperative ECoG, particularly in the context of epilepsy surgery, has shown higher rates of complications such as infection and hemorrhage.^33,34^ These findings, however, are based on subdural grid placements performed via craniotomy, which are significantly more invasive than the burr hole–based approach used in DBS. As such, the favorable safety profile demonstrated here cannot be directly translated to long-term or implantable ECoG systems without further investigation.

### High-Resolution, Multi-Subject Data Integration

This report presents anatomical and neurophysiological data obtained during awake DBS procedures using high-density ECoG strips, analyzed at both single and multiple-subject levels. A key advantage of this approach is the ability to perform anatomically standardized, group-level analysis with high spatial resolution. Electrode contacts were localized in ACPC space and transformed into MNI coordinates, enabling accurate alignment of data across subjects on a common brain surface. In this cohort, 21 ECoG strips, each containing 63 contacts were successfully registered to a single MNI brain (Figure 4), yielding a total of 1,323 electrode positions. This allowed for broad cortical surface coverage and high-resolution visualization of physiological activity across patients.

The combination of high-density electrode arrays and a standardized anatomical analysis pipeline represents a methodological advancement in intraoperative ECoG research. This approach enables investigation of cortical activity during both resting state and intraoperative tasks, with significantly improved spatial precision compared to single-subject analyses. As the number of subjects increases, the analysis gains further resolution and reproducibility, supporting its use in larger-scale studies of movement disorder physiology.

New research continues to emerge in the field of electrocorticography, especially with regard to the advancements in the development of closed-loop stimulation systems for movement disorders.^20–23^ High-resolution neurophysiological analysis by multiple subject analysis is a promising advancement of the well-established tool of electrocorticography. It provides a practical and standardized approach for collecting and combining high-resolution cortical data.

### Research opportunities and potential clinical applications

ECoG during DBS procedures represents a unique opportunity for neurophysiological research of movement disorders^35^ ^36^ ^37^ ^38^ ^39^. The burr hole created for the DBS lead is the entrance point through which the ECoG strip can be placed. Cortical activity recording can be obtained from the circumferential cortical surface area surrounding the burr hole, providing a large area for neurophysiological data collection (Figure 5). The area of recording depends on the movement disorder studied and the site of the burr hole^40^. A large number of papers have been published in the context of understanding the neurophysiology of movement disorders by incorporating ECoG recordings^41^ ^42^.

### Limitations

ECoG during DBS is a useful research technique given the limited opportunities to record human neural activity directly. Stimulation can be performed, and recordings can be obtained from subcortical targets and the cortical areas surrounding the burr hole. However, some parts of the cortex are more challenging to access from the standard burr hole locations, such as the medial prefrontal cortex and orbitofrontal cortex. Additionally, as these studies are performed intraoperatively, there is limited surgical time designated for research, and the recordings and the performance of the subject may be influenced by anesthetics, narcotic analgesics, and the surgery itself. This may be addressed by recent advancements in implantable ECoG-based closed-loop stimulation systems, which provide real-time recordings in a less artificial environment.

### Conclusion

This report demonstrates that high-density electrocorticography during awake DBS surgery is a safe, technically feasible method for obtaining high-resolution cortical data. By integrating anatomical localization across subjects using standardized coordinate systems, this approach enables group-level analyses with improved spatial resolution. These findings support the use of intraoperative ECoG for advancing movement disorder research and provide a foundation for future closed-loop neuromodulation research.

## Data Availability

All data produced in the present study are available upon reasonable request to the authors

## Acknowledgement

The authors express sincere gratitude to our participants, the neurosurgery department, and the depatmmet of neurology, Neuromodulation and Advanced Treatments Center.

## Conflict of Interest Statement

WSA serves on the advisory board of Longeviti NeuroSolutions. He also serves as a compensated consultant to Globus Medical and iota Biosciences. The other authors declare no competing financial interests or conflicts of interest.

## Notes

**Conflict of Interest:** The authors declare no competing financial interests.

### Competing Interest Statement

The authors have declared no competing interest.

### Funding Statement

This study did not receive any funding

### Author Declarations

The Johns Hopkins Institutional Review Board approved the study.

